# Brain Fluidity as a Functional Marker of Tau-Related Neurodegeneration in Alzheimer’s Disease

**DOI:** 10.1101/2025.07.04.25330744

**Authors:** Camille Mazzara, Giuditta Gambino, Gian Marco Duma, Matteo Neri, Giuseppe Giglia, Tommaso Piccoli, Fabrizio Guajana, Albert Comelli, Antonino Tuttolomondo, Michele Migliore, Pierpaolo Sorrentino

## Abstract

Alzheimer’s Disease (AD) is the most common form of dementia and one of the leading neurodegenerative disorders worldwide. Neurophysiopathologically, AD is characterized by neuronal death, accompanied by the alteration of tau protein in the cerebrospinal fluid (CSF) and beta-amyloid plaques in brain tissue. Although significant progress has been made in identifying the biological markers of AD, a deeper understanding of how these changes lead to functional disruptions in brain dynamics - and ultimately to clinical symptoms - remains crucial. In this context, quantitative electroencephalography (qEEG) has emerged as a valuable neurophysiological tool for studying the temporal dynamics of large-scale brain activities with high temporal resolution. A growing body of research has recently focused on brain fluidity, a metric of the brain’s ability to dynamically reconfigure its functional connectivity patterns over time - reflecting its capacity for adaptive processing and cognitive flexibility. In Alzheimer’s disease, this fluidity may be altered, suggesting a reduced ability of the brain to shift between different functional states in response to internal or external stimuli. In this study, we explored alterations in brain fluidity in AD by analyzing EEG data from 28 patients with a clinical diagnosis of Alzheimer’s disease and 29 age-matched healthy controls. Following preprocessing and source reconstruction, we computed functional connectivity using the Phase Locking Value (PLV) within sliding time windows. Brain fluidity was then quantified as the average similarity between successive PLV connectivity matrices, for each canonical frequency band. Our findings reveal frequency-specific alterations of fluidity in AD patients, particularly in the theta and beta frequency bands. Moreover, we found that reduced fluidity was directly correlated with lower scores on the Mini-Mental State Examination and with higher levels of CSF biomarkers, such as the levels of total tau protein and those of phosphorylated tau. These results suggest that decreased brain fluidity may serve as a functional marker of tau-related neurodegeneration and cognitive impairment in AD. In conclusion, our study provides evidence that the temporal variability of functional connectivity, captured through EEG-based brain fluidity, represents a promising functional biomarker, potentially aiding in early diagnosis and monitoring of disease progression.

## Introduction

Alzheimer’s Disease (AD) is the most common form of dementia and ranks among the foremost neurodegenerative diseases worldwide (Mielke, 2018; Uddin et al., 2018). Characterized by a progressive decline in cognitive and functional abilities, AD eventually impairs nearly all aspects of daily living (Giebel et al., 2014; Maresova et al., 2020; Tekin et al., 2001). Considering its complex, multifactorial nature, a comprehensive understanding of AD development and progression requires an integrated perspective that bridges alterations in brain structure, changes in neural dynamics, and the resulting cognitive decline (Hampel et al., 2018; Ibanez et al., 2024) (fig.1). Previous literature highlights that AD is characterized by changes in brain structure, notably area-specific brain atrophy and neuronal death (de Paula et al., 2009; Duan et al., 2012; Serrano-Pozo et al., 2011). According to current diagnostic guidelines, AD is associated with: 1) reduced Aβ42 in cerebrospinal fluid (CSF) and increased amyloid levels in the brain, as revealed by amyloid positron emission tomography (PET) mapping and 2) increased phospho-tau in both CSF and tau PET mapping (Jack Jr. et al., 2024).

**Fig. 1.**
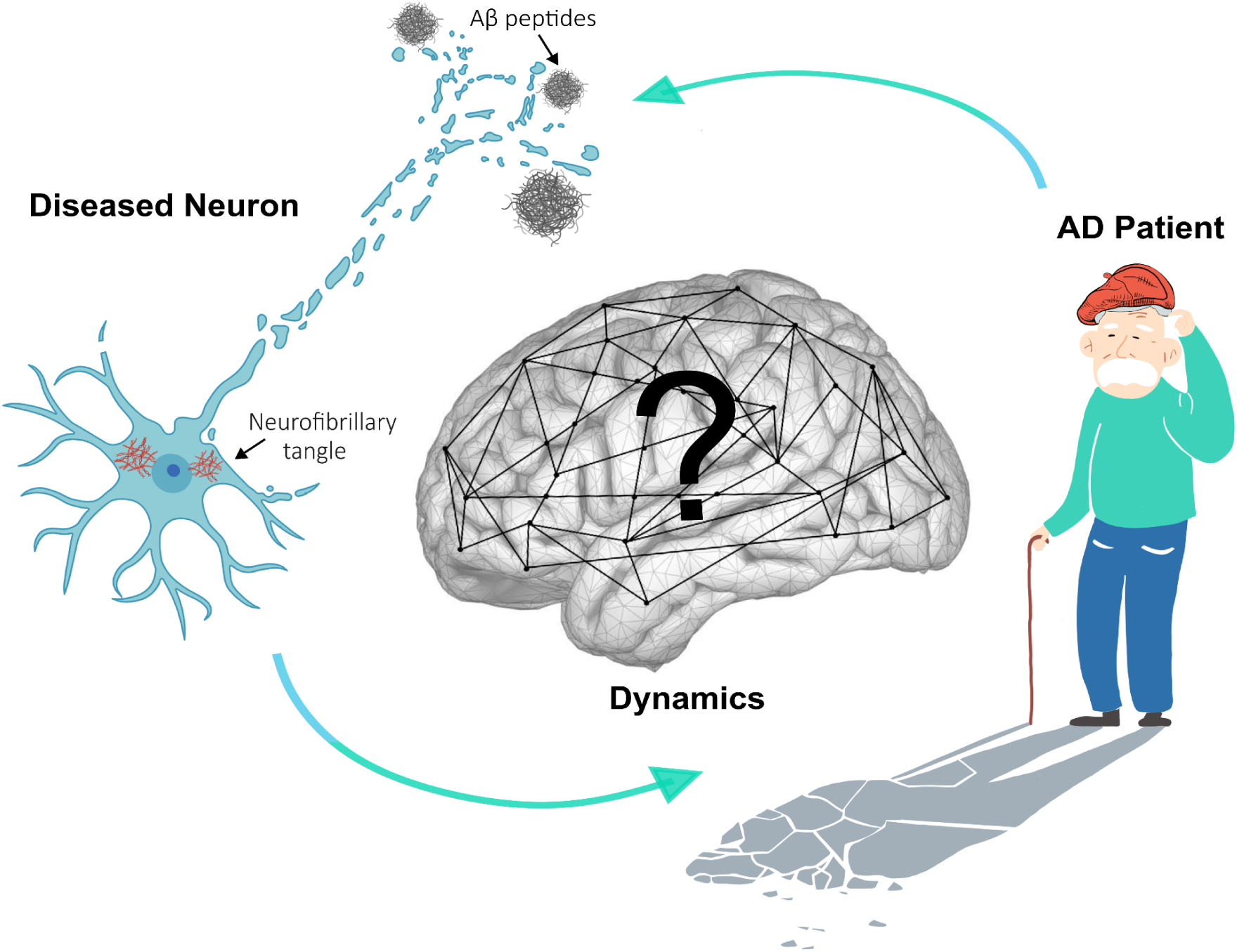
Conceptual framework of the study. Alzheimer’s disease (AD) is characterized by progressive neuronal death, illustrated to the left by a degenerating neuron along with its key biomarkers: amyloid-beta and tau proteins. On the right, a symbolic representation of cognitive decline is shown as a confused elderly patient whose shadow is dissolving, highlighting the impact of AD symptoms on identity and function. While neuronal degeneration and clinical manifestations are well-established in AD, the effects on large-scale brain dynamics remain poorly understood. The central panel depicts the brain as a dynamic network, whose flexible functional configurations may be compromised in AD. This study aims to investigate whether neuronal death leads to brain networks with different dynamics. Understanding these network-level changes could offer new insights into AD pathophysiology.

Structures within the temporal lobe, such as the hippocampal subfields (CA1,CA2,CA3) and the entorhinal cortex, are particularly vulnerable to damage in AD (Bierer et al., 1995; Fukutani et al., 2000). Due to significant atrophy observed in these areas, medial temporal lobe shrinkage is often used as a diagnostic marker for neurodegeneration (Elsiddig et al., 2018; Kehoe et al., 2014), as revealed by fluorodeoxyglucose PET (FDG-PET), total tau in CSF, and magnetic resonance imaging (MRI) of brain atrophy in the temporoparietal cortex and the hippocampi (Jack Jr. et al., 2024). Nonetheless, among cost-effective and non-invasive diagnostics measures, electroencephalography (EEG) could hint for synaptic dysfunction and individual altered neurophysiological traits (Jack Jr. et al., 2024).

AD symptoms primarily involve memory loss and cognitive decline, impacting language, problem-solving, and spatial awareness (Buckley et al., 2015; Jahn, 2013; Morganti et al., 2013; Verma and Howard, 2012). Disorientation in space and time, difficulty in recognizing familiar people and objects, and impairments in executive functions are also common (Allain et al., 2013; Belleville, 2010; Monacelli et al., 2003; Peters-Founshtein et al., 2024). As the disease progresses, patients may experience mood and behavioral changes, including apathy, anxiety, and irritability, causing significant impairments in daily functioning and independence (Botto et al., 2022; Cummings et al., 2006; Mega et al., 1996; Zhao et al., 2016).

Cognitive symptoms affect higher functions. In contrast to simpler functions, which localize more to specific regions, higher cognitive functions likely arise from the coordinated interactions among multiple brain areas (Romano et al., 2023). Following this reasoning, a long standing hypothesis in the field holds that AD disrupts brain functions by altering communications among distributed brain networks. Along these lines, AD has been understood as a disconnection syndrome (Delbeuck et al., 2007; Paitel et al., 2025). Consequently, extensive research adopted holistic approaches to investigate brain function, focusing on the description of the organization of activities over the large scale (Palop and Mucke, 2016). In this context, theoretical tools from network science and signal processing have been employed to describe interactions among brain regions across a broad range of different temporal and spatial scales. These approaches typically investigate interactions by analysing statistical dependencies between brain signals (Friston, 2011; Stam, 2014).

However, a clear pattern of disease-related alterations did not emerge across studies (Paitel et al., 2025). One possible reason may be that traditional network approaches disregard the dynamics of the interactions. For instance, Functional Connectivity (FC) measures statistical dependencies between signals from different brain regions over a given time interval, as a proxy of synchronized or coordinated activities (Fornito et al., 2015; Friston, 2011). Hence, FC provides a static picture, capturing average connection strengths over time, based on the assumption that brain networks remain relatively static in the resting brain (Hutchison et al., 2013). However, complex cognitive processes require the dynamic recruitment of different brain areas to respond to changing environmental demands. Such complex dynamics manifest themselves as continuous adjustments in connectivity (Zalesky et al., 2014). As such, a healthy brain generates complex dynamics, which might be lost in disease. Hence, the quantification of the “flexibility” of the network dynamics might be linked to neurodegenerative processes and allow a deeper understanding of the network mechanisms underpinning AD (Cipriano et al., 2024; Polverino et al., 2022; Sorrentino et al., 2021).

To quantify to what extent brain dynamics are flexible, different works focused on studying the Functional Connectivity Dynamics (FCD) (Preti et al., 2017). FCD accommodates the non-stationary nature of brain networks, marked by rapid shifts between distinct connectivity states (Hansen et al., 2015). Rather than focusing on a single “snapshot” of connectivity, FCD analyzes fluctuations in connectivity patterns over time intervals, unveiling the dynamic nature of interactions between brain regions, and capturing the brain’s inherent flexibility and adaptability. These fluctuations might stem from complex non-linear interactions among brain regions, which generate synchronized activities, itinerant dynamics, and multistability (Heitmann and Breakspear, 2018). It has been seen that the statistical characteristics of the dynamics span from a strictly “order-driven” dynamic, where the average FC remains stable, to a purely “random-driven” scenario, where FC fluctuations are entirely uncorrelated over time (Battaglia et al., 2020). This framework would place the “optimal” dynamics in between these two extremes.

More recently, a new metric called *fluidity* has been introduced to capture the richness and adaptability of large-scale brain dynamics. Alterations in fluidity — whether excessive or diminished— have been reported across various neurological diseases (Battaglia et al., 2020; Breyton et al., 2024; Yalçınkaya et al., 2023). Specifically, reduced fluidity may reflect rigid and stereotyped connectivity patterns, limiting adaptability. Conversely, excessive fluidity might reflect a lack of coordinated activity. We hypothesize that in AD brain dynamics deviate from the optimal regime, resulting in suboptimal dynamics.

Notably, at symptom onset, AD patients show a general slowing of brain rhythms (Jafari et al., 2020; Jeong, 2004). In terms of brain oscillations, this is mirrored by a decrease in spectral power in higher frequency bands (beta/gamma), in favor of a more preponderant increase in lower ones (theta/delta) (Jafari et al., 2020; Jeong, 2004). Importantly, this high-to-low-frequency ratio is significantly more decreased in clinically-diagnosed AD patients compared to healthy subjects (Jafari et al., 2020; Jeong, 2004). This frequency-specific reorganization suggests that changes in fluidity are not uniform but instead vary across frequency bands, potentially correlating with distinct cognitive deficits.

Building on this literature we expect to find changes that are frequency-band dependent, potentially exhibiting opposite trends between higher and lower frequencies. These frequency-specific changes in network dynamics—such as alterations in properties like fluidity—may index the impact of neuronal death on higher-order cognitive functions (Fig. 1). Therefore, we propose that fluidity alterations in AD are both frequency-dependent and correlated with disease markers of neuronal death such as CSF levels of Aβ42, p-tau, and t-tau, as well as cognitive scores.

In particular, we sought to determine whether fluidity could serve as a predictive marker. This hypothesis is grounded in the premise that deviations in the temporal variability of connectivity may reflect underlying disease processes.

To test this hypothesis, we analyzed source-reconstructed EEG data, leveraging its optimal temporal resolution, from a cohort of 28 AD patients and 29 age-matched healthy controls. All patients underwent lumbar puncture to quantify CSF biomarkers (Aβ42, p-tau, and t-tau), and cognitive performance was assessed using the Mini-Mental State Examination (MMSE). For each canonical frequency band, we characterized brain dynamics from the EEG data in terms of fluidity. If our hypothesis holds, we expect to observe frequency-specific deviations in fluidity, either reduced or excessive, in AD patients compared to controls. Furthermore, these deviations should correlate with the burden of pathological markers and the degree of cognitive impairment, supporting the potential of fluidity as a novel marker of disease-related brain dysfunction.

## Results

To investigate the role of fluidity in AD, we analyzed EEG data from AD patients and healthy controls. Our aim is to explore whether elements of brain dynamics correlate with the presence of pathological markers and clinical disability. By doing so, we sought to determine whether fluidity could serve as a predictive marker. This hypothesis is grounded in the premise that deviations in the temporal variability of connectivity may reflect underlying disease processes. The analysis pipeline is shown in Fig.2. The analysis began with the preprocessing of EEG data for both AD patients and Controls, which involved filtering and cleaning the signals to remove artifacts. Following this, source reconstruction was conducted using a default anatomy as follows: This process involved solving the inverse problem to estimate the neural activity originating from the brain’s surface, based on the recorded electrical potentials. The Desikan-Killiany atlas was utilized to parcellate the brain into 68 distinct regions of interest. To gain insights in the temporal scales, neural activities from each region were subsequently analyzed to extract signals in the canonical frequency bands: theta (4-8 Hz), alpha (8-14 Hz), beta (14-30 Hz), and gamma (30-40 Hz). To assess connectivity, we calculated the Phase Locking Value (PLV), a widely used measure of phase synchronization between pairs of brain regions (Lachaux et al., 1999). A high PLV indicates strong phase coherence, suggesting robust communication, while a low PLV denotes weaker synchronization and potentially less effective interaction. The PLV was computed in sliding windows, capturing connectivity changes over time. Subsequently, Pearson’s correlation was computed across the PLV matrices from consecutive windows, obtaining a dynamic functional connectivity (dFC) matrix, which reflects the temporal evolution of inter-regional synchronization. By analyzing the variance of PLV across these sliding windows, the temporal variability of functional connectivity was quantified. This variance provides a measure of flexibility, offering insights into how the brain’s connectivity adapts dynamically.

**Fig. 2.**
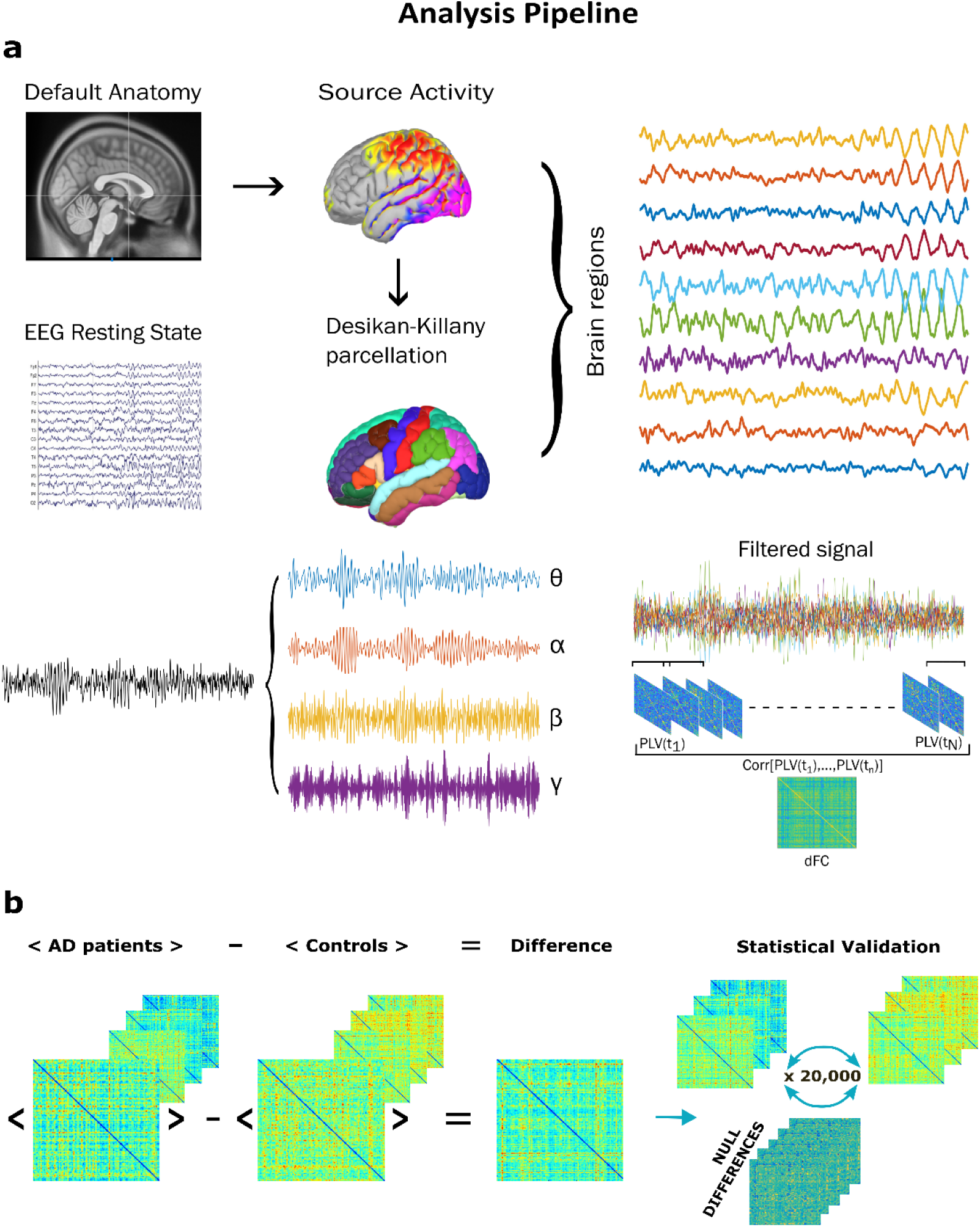
a. Overview of the analysis pipeline. EEG signals from AD patients and healthy controls underwent preprocessing and source reconstruction using a Default Anatomy. Brain activities were reconstructed in 68 regions using the Desikan-Killiany atlas and filtered into canonical frequency bands. Functional connectivity dynamics were assessed via the Phase Locking Value (PLV) computed over sliding windows, capturing time-resolved phase synchronization across regions. The variance of these dynamic connectivity patterns was quantified as a proxy for network fluidity; **b.** Group-level analysis was conducted to assess whether the overall average fluidity differed significantly between AD patients and healthy controls. For each subject, we computed the mean value of the time-by-time matrix, and then compared the group averages between conditions. To validate the observed difference, we applied a non-parametric permutation test. The group labels (i.e., AD or control) were randomly shuffled across subjects 20,000 times. At each iteration, the absolute difference in group means was recomputed. This procedure generated a null distribution of differences under the assumption that there is no difference in the dynamics across groups.

We assume that communication between two brain areas occurs at the same frequency band and involves a synchronization mechanism. The analysis of fluidity was performed for each frequency band (theta, alpha, beta, and gamma) across all participants (Fig.3a). Significant differences were observed in the theta and beta bands between Alzheimer’s disease (AD) patients and healthy controls. Specifically, in the theta band (4–8 Hz), fluidity was significantly higher in AD patients as compared to controls (p = 0.0018). Conversely, in the beta band (14–30 Hz), fluidity was significantly lower in AD patients compared to controls (p=0.0006). In Fig. 3b, the PLV matrices averaged across all patients are shown for the AD group (to the left) and the control group (to the right), in the theta band (top), and in the beta band (bottom). As shown, when the correlation is higher, PLV matrices are more similar across time, meaning that there are fewer different states. In this case, the PLV matrices appear more stable and less variable, as if the brain is “stuck” in certain stable networks and struggles to transition out of them. This suggests that the system is more rigid, with reduced flexibility in exploring different states. However, the effect differs across frequency bands, aligning with our initial frequency-dependence hypothesis. This trend suggests a redistribution of dynamic functional connectivity, with AD patients favoring increased flexibility for slow activities and more stereotyped, faster dynamics.

**Fig. 3.**
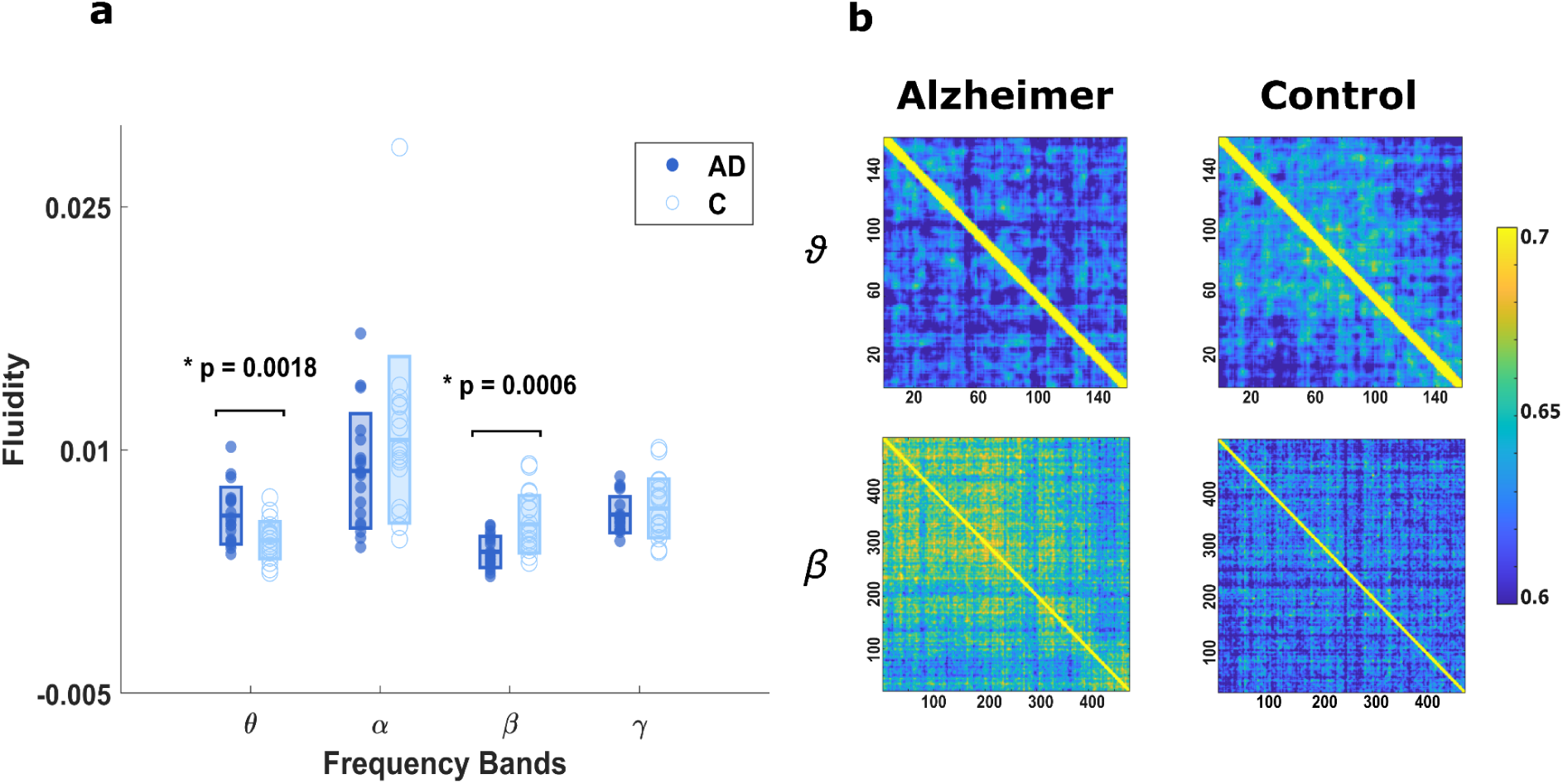
Group differences in fluidity across frequency bands. **a.** Distribution of individual fluidity values in AD patients and controls across theta (4–8 Hz), alpha (8–14 Hz), beta (14–30 Hz), and gamma (30–40 Hz) bands. AD patients showed significantly higher fluidity in the theta band (p = 0.0018) and significantly lower fluidity in the beta band (p = 0.006) as compared to controls; **b.** Group-averaged Fluidity matrices for the theta (top) and beta bands (bottom), shown for AD patients (left) and controls (right). Connectivity matrices in AD appear more similar across time in the beta band (stronger Correlation), indicating lower variability and reduced network flexibility. Conversely, theta-band dynamics are more variable in AD, suggesting altered distribution of flexibility across frequencies.

We found that the differences in dynamic flexibility between AD patients and Controls are selectively in the theta and beta frequency bands. Specifically, in the theta band, we observed hyper-fluidity in AD patients, whereas in the beta band, we found hypo-fluidity. To interpret these findings, we explored their association with key biomarkers, total tau (tTau), phosphorylated tau (pTau), and amyloid-beta (Aβ), as well as with clinical measures such as the Mini-Mental State Examination (MMSE) score. We then calculated the correlations between the dynamic flexibility in the theta and beta bands and these biomarkers. Interestingly, no significant correlations were found between theta fluidity and the clinical measures. However, for beta fluidity, we found a significant negative correlation with tTau and pTau (ρ= -0.51, p = 0.03 and ρ= -0.56, p = 0.01, respectively), while no significant correlations were observed with Aβ levels. From a clinical perspective, these results suggest that the fluidity, or the ability of the system to operate in a metastable dynamic, particularly for faster activities like beta and gamma (for gamma, see Figure S1), is more closely linked to cellular damage, as indicated by increased tau protein levels in the cerebrospinal fluid. This could imply that changes in the ability of the brain to flexibly switch between different dynamic states is associated with neurodegenerative processes, specifically tau-related damage.

Using a multilinear regression model, we aimed to predict clinical disability, as measured by the Mini-Mental State Examination (MMSE), incorporating variables such as age, sex, total tau (tTau) levels, and beta fluidity as predictors. This method allows us to assess the individual contribution of each predictor while accounting for the effects of the others.

Adding beta fluidity to the model significantly enhanced its predictive power, as demonstrated in Figure 4. The final model explained 49% of the variance in MMSE scores (R² = 0.49; Adjusted R² = 0.34) when all four predictors were included. The individual contributions of the predictors were as follows: age (β =2.0829, ρ = 0.0037), tTau (β = -0.4973, ρ = 0.0286), sex (β = -0.0956, ρ = 0.4990),, and fluidity in beta band (β = -0.4868, ρ = 0.0232). These results suggest that beta fluidity provides additional explanatory power for clinical outcomes when combined with other variables. To ensure the robustness of our findings, we validated the model using a leave-one-out cross-validation (LOOCV) approach. In LOOCV, one subject is left out for testing while the model is trained on the remaining individuals, and this process is repeated for each participant. This method reduces the risk of overfitting and provides an unbiased estimate of the model’s performance on new, unseen data.

**Fig. 4.**
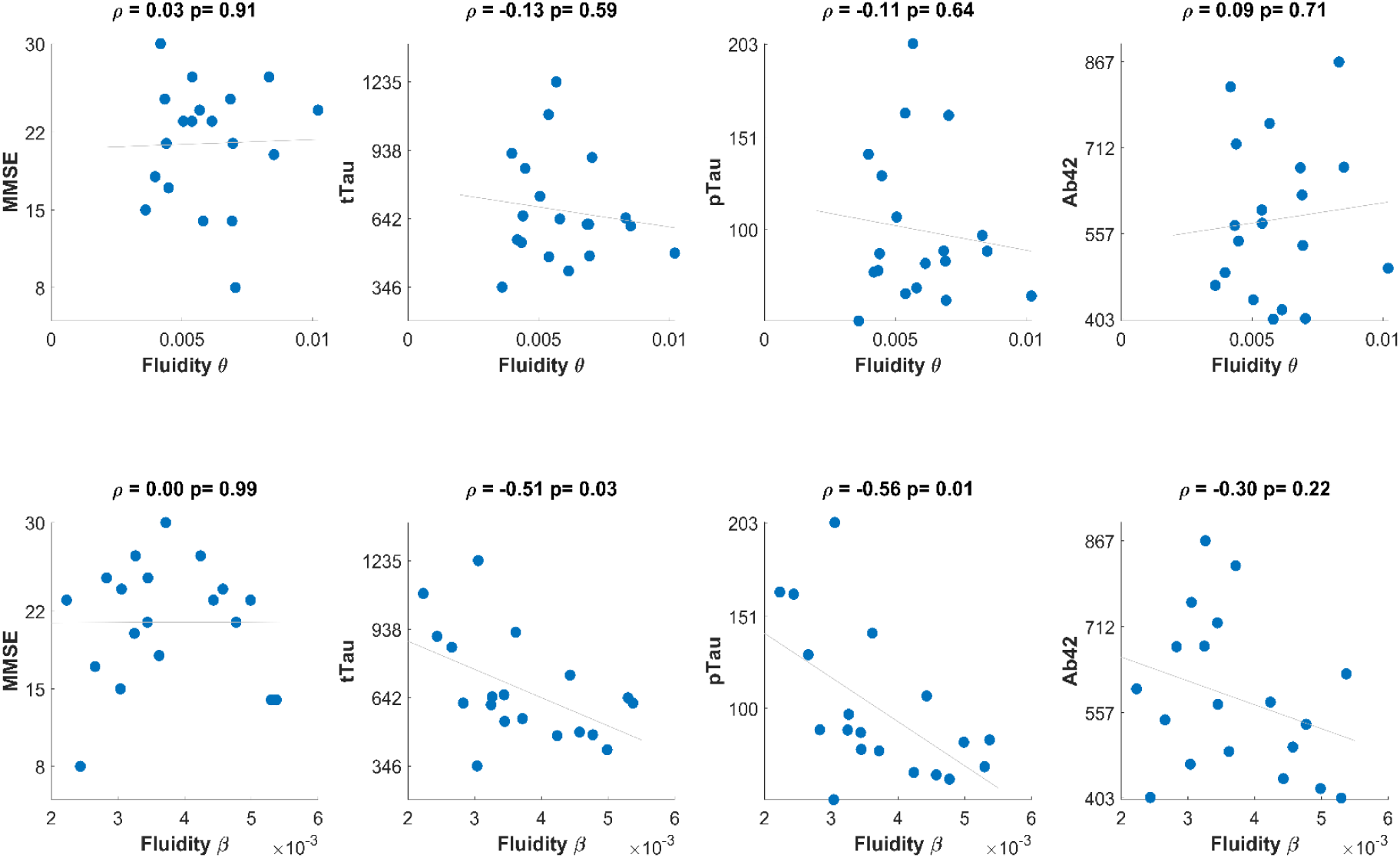
Associations between fluidity and clinical/biomarker measures in Alzheimer’s disease. *(Top row)*: Correlations between theta-band fluidity and MMSE score, total tau (tTau), phosphorylated tau (pTau), and amyloid-beta 42 (Aβ42) levels. No significant correlations were found in the theta band. *(Bottom row)*: Correlations between beta-band fluidity and the same clinical and biomarker measures. Significant negative correlations were observed between beta fluidity and both tTau (ρ= -0.51 , p = 0.03) and pTau (ρ= -0.56, p = 0.01), but not with MMSE or Aβ42. These results suggest that beta-band fluidity may reflect tau-related neurodegenerative processes more closely than cognitive status or amyloid burden.

The cross-validated model produced similar results, with an explained variance of 49% (R² = 0.49; Adjusted R² = 0.34). Predictor contributions in the cross-validated model were age (β =2.03, ρ = 0.0073), tTau (β = -0.7017, ρ = 0.0387), sex (β = -0.1780, ρ = 0.5290), and fluidity in beta band(β = -0.6117, ρ = 0.0340). Fig.5 illustrates the predicted versus observed MMSE scores (panels b and e) and an evaluation of residuals (panels c and f), demonstrating good alignment between the model’s predictions and empirical data. The increase in explained variance highlights the clinical relevance of beta fluidity as a meaningful correlate of cognitive disability in Alzheimer’s disease.

**Fig. 5.**
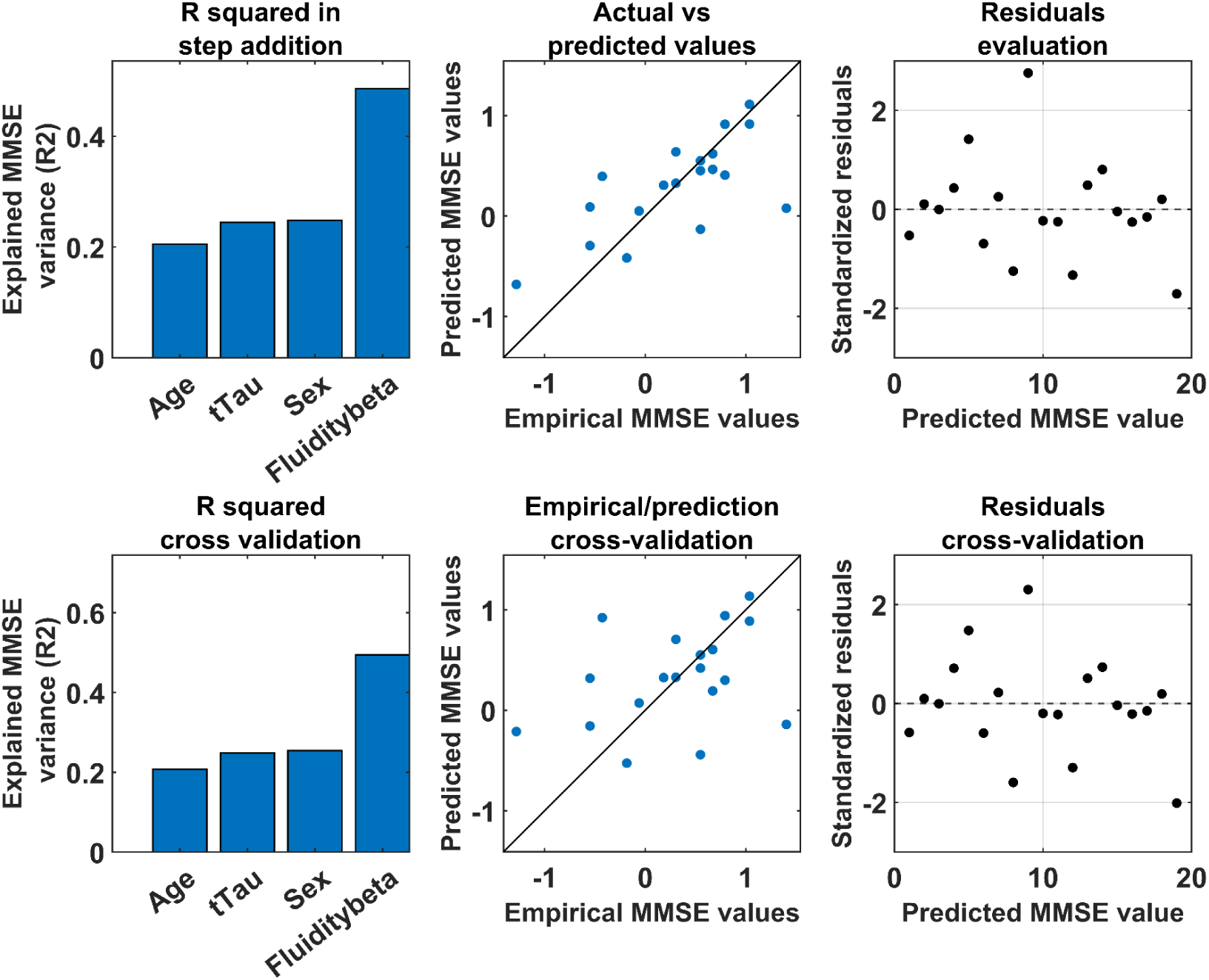
Multilinear regression model predicting MMSE scores in Alzheimer’s disease. ***(a,d)*** Explained MMSE variance using age, sex, total tau (tTau), and beta fluidity as predictors for the full model **(a)** and the leave-one-out cross-validation (LOOCV) model **(d)**; ***(b, e)*** Predicted versus observed MMSE scores for the full model **(b)** and the leave-one-out cross-validation (LOOCV) model **(e)**; ***(c, g)*** Residual plots for the same models.

To avoid multicollinearity, we excluded phosphorylated tau (pTau) from the model due to high Variance Inflation Factor (VIF) values exceeding the threshold for multicollinearity. The VIF quantifies how much the variance of a regression coefficient is inflated due to multicollinearity with other variables. High VIF values indicate that predictors are not independent, which can destabilize the model and make it difficult to accurately assess the contribution of each variable.

By excluding pTau, we ensured that the remaining predictors were sufficiently independent to produce reliable estimates.

At this point, since we found a correlation between global fluidity and tau (a marker of neuronal cell death), we asked whether the observed changes in fluidity could be linked to specific brain regions rather than being uniformly distributed. Despite using EEG, which has limited spatial resolution, we aimed to investigate whether certain regions contribute more to the observed changes in fluidity.

To address this, we sought to transition from a global metric of fluidity to a region-specific, or nodal, metric. Specifically, we wanted to interpret the role of each brain region in terms of how much it contributes to overall fluidity. Alternatively, this could also be conceptualized as how much each region constrains the brain’s ability to maintain fluidity.

To quantify this, we recalculated global fluidity iteratively, removing one region at a time (Fig. 6a). For each iteration, we computed the difference between the original global fluidity and the fluidity after removing the specific region. This provided a measure of each region’s impact on global fluidity.

**Fig. 6.**
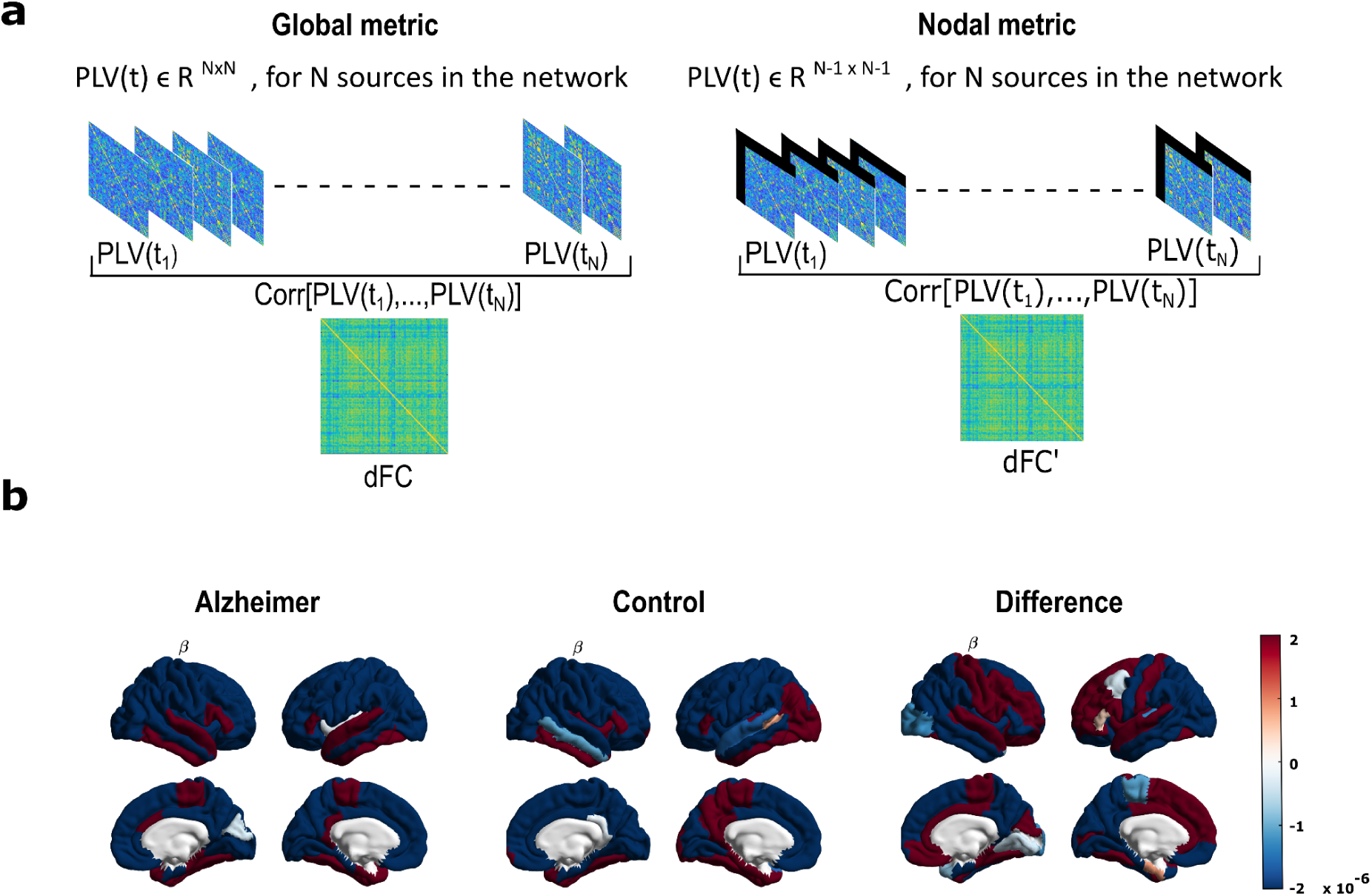
Regional contributions to global fluidity in Alzheimer’s disease. **a)** Schematic representation of dynamic fluidity computation: standard global fluidity estimation (left) and nodal contribution analysis by region-wise exclusion (indicated by black bar) (right); ***(b)*** Brain surface maps showing regional contributions to global fluidity in the beta band for Alzheimer’s disease (AD) patients (left), healthy controls (middle), and the difference between groups (right). Warmer colors indicate regions whose removal leads to a stronger reduction in global fluidity, implying higher contribution.

We then plotted these values on the brain for both Alzheimer’s disease (AD) patients and controls, as well as the differences between the two groups (Fig. 6b).

Our results showed that certain regions, particularly in the lower temporal lobe, contribute more to global fluidity in both AD patients and controls. These regions ( in red on the brain plots), once removed, caused a greater reduction in global fluidity. Interestingly, the temporal regions were important not only in AD patients but also in healthy controls. This suggests that these regions play a key role in fluidity across groups, and AD patients tend to reflect the same patterns observed in healthy individuals.

However, when examining the differences between AD patients and controls, no clear pattern emerged (Fig. 6b). The differential map represents the difference in how much each region’s removal affects global fluidity in the two groups. Positive values indicate that removing a region has a greater impact on global fluidity in controls than in AD patients, while negative values mean the opposite. For example, “red” regions in AD indicate that removing the region reduces fluidity more in AD patients, suggesting that these regions contribute more to fluidity in AD. Conversely, “blue” regions contribute less to fluidity in AD, perhaps because their function is already impaired, resulting in a smaller effect on fluidity when removed.

We also performed a permutation test by shuffling the labels of AD patients and controls to determine the statistical significance of our findings. Although two regions appeared significant, the significance did not hold after correction for multiple comparisons.

## Discussion

In this study we tested the hypothesis that large-scale brain dynamics are altered in Alzheimer’s disease (AD), with alterations being frequency-specific and correlating with both pathological biomarkers and cognitive impairment. To address this question, we adopted a dynamic approach by quantifying functional connectivity dynamics (FCD) using a novel metric called *fluidity.* This measure captures the brain’s ability to dynamically reorganize and adapt its activities over time, reflecting its underlying ability to support cognitive adaptability and efficient information processing (Battaglia et al., 2020).

Our results provide novel insights into the pathophysiology of AD, revealing a frequency-specific reorganization of network dynamics. Consistent with our hypothesis that AD disrupts the optimal balance of brain functional architecture, we observed a frequency-specific alteration in fluidity: AD patients showed significantly increased fluidity in the theta band (4–8 Hz) and reduced fluidity in the beta band (14–30 Hz) compared to healthy controls. This double dissociation supports the notion that AD pathology may differentially affect brain rhythms and their temporal organization, possibly reflecting the altered engagement of neural circuits in different cognitive operations (Palmigiano et al., 2017), suggesting a redistribution of dynamic flexibility across frequency bands.

The increased theta fluidity in AD patients may reflect an instability or loss of coordinated theta-band activities. Excessive fluctuations (with respect to the presumably optimal regime observed in healthy subjects) may reflect disorganized communication across brain regions. Theta oscillations are known to support memory and attention functions (Benchenane et al., 2011; Herweg et al., 2020), which are domains heavily impacted by AD. However, we did not observe a significant correlation between theta fluidity and either CSF biomarkers or cognitive scores (MMSE), suggesting that theta hyper-fluidity might not directly relate to pathological load or cognitive disability but may instead reflect compensatory or maladaptive dynamics. As structural integrity deteriorates, the brain may rely more heavily on slower oscillations to maintain functional communication.

By contrast, the decreased fluidity in the beta band was significantly and negatively associated with both total tau and phosphorylated tau proteins levels—two well-established markers of neuronal degeneration (Delbeuck et al., 2007; Watanabe et al., 2019; Yu et al., 2021). Furthermore, when included in a multiple regression model alongside age, sex, and tTau, beta fluidity significantly improved the prediction of MMSE scores. This finding highlights beta-band dynamics as particularly informative for understanding the relationship between network rigidity and cognitive decline.

Interestingly, decreased beta-band fluidity does not correlate with amyloid-beta (Aβ) levels. Although Aβ accumulation remains a diagnostic hallmark and is often used for diagnostic purposes (Hampel et al., 2021; Zhang et al., 2023), it lacks specificity for ongoing neurodegeneration (Dubois et al., 2016; Jansen et al., 2015; Kurkinen et al., 2023). The inverse correlation between beta fluidity and p/t tau levels in the CSF reinforces the notion that the loss of flexible dynamics is related to neuronal dysfunction and death. Thus, the observed relationship between reduced beta fluidity and elevated tau levels likely reflects the breakdown of coordinated, flexible neural activity due to tau-mediated structural and functional damage. Beta oscillations are critically involved in top-down cognitive control, attention, and sensorimotor integration (Engel and Fries, 2010; Stoll et al., 2016). A reduction in beta fluidity may therefore signal impaired adaptive regulation across cognitive domains, reflecting a network-level manifestation of impaired executive function. As such, it could serve as a sensitive marker of pathological processes underlying AD-related cognitive dysfunction.

Finally, the differences we observed are consistent with a growing body of literature indicating that neuronal loss in AD patients leads to a global shift in neurophysiological activity toward a slower regime compared to healthy individuals (Gallego-Rudolf et al., 2024). This shift is typically marked by reduced high-frequency activity, such as in the beta band, and increased low-frequency activity, particularly in the theta band. These alterations might be mirrored by the increase of network fluidity that we observed in the theta band and the decrease of fluidity in the beta band. In line with previous work, our findings highlight the presence of opposite patterns in low- and high-frequency bands as a hallmark of AD-related neurophysiological changes. Moreover, our results suggest that neuronal degeneration and cognitive decline in AD beyond being associated with changes in spectral power, can be related to broader network-level disruptions, more specifically, to a loss of fluidity in network dynamics in the beta band.

Our findings align with the conceptualization of AD as a disconnection syndrome (Delbeuck et al., 2007; Paitel et al., 2025), where cognitive decline arises not solely from regional atrophy but from disrupted interactions between brain regions. Prior studies using static functional connectivity (FC) measures have yielded mixed results (Paitel et al., 2025), perhaps due to their inability to capture the temporal variability of network interactions. Most EEG connectivity studies in AD to date rely on *static* measures (e.g., Pearson correlation, mutual information, coherence, phase-locking value), often overlooking the inherently dynamic nature of brain networks (Bastos and Schoffelen, 2015; Cao et al., 2022; Cohen, 2014). Moreover, only a minority of studies have implemented source reconstruction techniques, limiting spatial specificity and the interpretability of observed connectivity changes. This lack of consistency across studies is further compounded by methodological differences and variability in participant characteristics, such as age or cognitive reserve. While reduced alpha-band connectivity is a relatively consistent finding in AD (Babiloni et al., 2016; Engels et al., 2015), findings in other frequency bands are more heterogeneous. Some studies have reported increased connectivity in the theta and delta bands, particularly in patients with mild cognitive impairment (MCI) (Jacini et al., 2018) or early-stage AD (Dauwels et al., n.d.; Vecchio et al., 2014), whereas others report conflicting results even within the same frequency ranges (Jeong, 2004; Smailovic and Jelic, 2019) (Jeong, 2004; Smailovic et al., 2019).

These discrepancies likely stem from the widespread use of static connectivity metrics, which are ill-suited to capture the moment-to-moment fluctuations that characterize healthy brain dynamics. A shift toward methods that reflect the temporal dynamics of neural activity may improve the replicability and interpretability of connectivity findings in AD.

By adopting a dynamic framework, we were able to characterize the temporal flexibility of neural communication, thereby capturing subtler alterations that are not evident in static approaches. This perspective is further supported by the fluidity framework, which extends beyond average connectivity and considers how rapidly and richly the brain switches between different connectivity states. Such transitions are fundamental to healthy cognitive function (Cipriano et al., 2024; Liparoti et al., 2024; Polverino et al., 2022; Sorrentino et al., 2021), particularly in adapting to changing environmental demands. Theoretical arguments support the notion that fluidity may reflect loss of computational capacity and diminished adaptability of the AD brain (Palmigiano et al., 2017).

Studies have demonstrated the importance of the frontal lobe in fluid intelligence (Duncan, 2005; Duncan et al., 1995). More specifically, functional Magnetic Resonance Imaging (fMRI) studies involving the RPM task in adults have demonstrated that a region in the anterior prefrontal cortex, known as the rostrolateral prefrontal cortex (RLPFC), is activated when participants engage in relational integration during RPM tasks (Christoff et al., 2003; Lee et al., 2006). We also explored the spatial specificity of fluidity by assessing each brain region’s contribution to the global dynamic profile. Temporal regions, especially in the lower temporal lobe, consistently emerged as major contributors in both AD patients and healthy controls. However, group differences in regional fluidity contributions were not statistically robust, perhaps due to the limited spatial resolution of EEG.

From a clinical standpoint, beta-band fluidity significantly enhanced the predictive accuracy of models for cognitive performance. These findings suggest that dynamic connectivity metrics may serve as sensitive and complementary biomarkers of disease progression, alongside traditional structural and molecular indicators, and could eventually support personalized monitoring and therapeutic strategies. Nonetheless, given the cross-sectional nature of our data, the ability of beta fluidity to predict individual outcomes must be validated in longitudinal studies.

In conclusion, our results underscore the utility of EEG-derived dynamic measures, specifically fluidity, as a functional readout of brain network adaptability in AD. The frequency-specific alterations observed in theta and beta bands mirror a pathological reorganization of connectivity dynamics, with beta fluidity, in particular, showing strong associations with tau pathology and cognitive impairment. These findings support the conceptualization of AD as a disconnection syndrome and highlight the promise of dynamic EEG biomarkers in advancing our understanding of disease mechanisms and improving clinical assessment and disease monitoring in neurodegenerative disorders. In fact, fluidity may emerge as a cost-effective and non-invasive biomarker, compared to current standards such as CSF assessment via lumbar puncture, which is invasive, resource-intensive, and often not well-tolerated by patients (Rueda et al., 2015). In contrast, EEG stands out as safe, accessible, and promising in detecting early electrophysiological changes in AD (Cacciamani et al., 2021). Unlike classical qEEG indices (e.g., band power or coherence), the concept of fluidity will capture the adaptability and complexity of large-scale network dynamics, potentially offering a more integrative and sensitive readout of disease-related dysfunctions. As such, fluidity may contribute to a new generation of EEG-based markers with both diagnostic and prognostic utility in clinical and preclinical stages of AD.

## Limitations

In this study, EEG source connectivity was computed using 19 scalp electrodes. While it is well known that using a low number of electrodes reduces the spatial accuracy of the results, previous studies have shown that meaningful information can still be extracted, even with limited channel counts. In fact, several investigations using 19, 32, or 64 electrodes have successfully examined large-scale brain networks under similar experimental conditions (Canuet et al., 2012; Hata et al., 2016; Kabbara et al., 2018; Vecchio et al., 2014). This is likely because their focus was, similarly to ours, on comparing global network organization between groups, rather than on resolving fine-grained, local network features. Notwithstanding, the fact that our analyses yielded significant correlations with clinical and pathological markers even with this limitation underscores the robustness and translational potential of the fluidity metric. This suggests that fluidity can be reliably assessed in standard clinical settings, broadening its applicability to a wider range of outpatient clinics and laboratories that may lack access to high-resolution EEG systems. To further validate our findings, we also performed the same analysis at the sensor level and observed similar trends to those obtained after source localization. This consistency supports the robustness of the reported effects, even with a relatively low electrode count. The corresponding results are provided in the Supplementary Materials (Fig.S2).

## Methods

### Participants

Participants in the study were recruited from the Neurology Unit, Department of Biomedicine, Neuroscience and Advanced Diagnostics (Bi.N.D. – University of Palermo, Palermo, Italy). All participants underwent a comprehensive general and neurological medical history assessment in the presence of a caregiver, a neurological physical examination, cognitive screening using the Mini-Mental State Examination (MMSE), and an evaluation of both basic (ADL) and instrumental (IADL) activities of daily living. The Mini-Mental State Examination (MMSE) was used as a measure of global cognitive functioning (Folstein et al., 1975). This test is widely applied to assess the overall cognitive status of individuals with Alzheimer’s disease and to monitor the severity and progression of cognitive decline (Ismail et al., 2010). The MMSE score ranges from 0 to 30, with lower scores indicating more severe cognitive impairment. At the time of the initial assessment, all participants also underwent a resting-state EEG recording, as described in the next section. The study protocol also included a lumbar puncture to identify individuals with a biological diagnosis of Alzheimer’s disease and to correlate CSF biomarker levels with previously recorded EEG data. All participants provided written informed consent, and all procedures were conducted in accordance with the Declaration of Helsinki and its subsequent amendments. Inclusion criteria for patients in the early stages of Alzheimer’s Disease (AD) were: Diagnosis of Alzheimer’s Disease dementia according to NINCDS-ADRDA and AT(N) criteria, Diagnosis of MCI due to AD according to the 2011 NIA-AA criteria and AT(N) framework, Corrected MMSE score > 17.5 Inclusion criteria for healthy controls (HC) were: Age > 50 years, Normal cognitive functioning with an MMSE score > 28, Preserved autonomy in activities of daily living, Absence of subjective cognitive decline (SCD) Exclusion criteria for all participants included: Pre-existing psychiatric disorders or other neurological diseases, Concurrent use of psychotropic drugs, cholinesterase inhibitors, or NMDA antagonists, Uncompensated endocrine disorders (e.g., hypo-/hyperthyroidism, Cushing’s syndrome), Decompensated liver cirrhosis or other hepatic diseases An FDG-PET pattern consistent with AD was defined as the presence of a spatial pattern of hypometabolism in regions typically affected by AD (hippocampus, medial and lateral temporal cortex, superior and inferior parietal lobules, posterior cingulate cortex, and precuneus) (Nobili et al., 2018). A CSF pattern consistent with AD was defined by low levels of Aβ42 or Aβ42/40 ratio, in combination with elevated pTau levels (Albert et al., 2011).

### EEG data recording

The EEG recordings lasted at least 10 minutes. Participants were instructed to keep their eyes closed for 5 minutes and then open for the remaining 5 minutes, while maintaining a relaxed wakeful state. Cortical electrical activity was recorded using 21 electrodes placed according to the international 10–20 system (i.e., Fp1, F3, C3, P3, O1, F7, T3, T5, Fpz, Fz, Cz, Pz, Oz, Fp2, F4, C4, P4, O2, F8, T4, T6), with a physical reference between Cz and Oz. The EEG signal was band-pass filtered (lower cutoff: 1.5 Hz; upper cutoff: 40 Hz) and sampled at 256 Hz. Horizontal and vertical eye movements were also recorded (frequency band: 0.3–70 Hz). Electrode impedance was kept below 5 kΩ. None of the patients were receiving treatment with acetylcholinesterase inhibitors or NMDA receptor antagonists at the time of EEG acquisition.

### EEG Preprocessing

19 standard scalp electrodes were considered, excluding the reference channels. EEG data were processed offline using EEGLAB (v.14.2.2b.22). The EEG signals were initially downsampled to 256 Hz, and successively band-pass filtered (2–45 Hz) using a finite impulse response (FIR) filter with a Hamming window. A standardized preprocessing pipeline was implemented to enhance reproducibility. For the present study, only the eyes-closed (EC) condition was analyzed, as it is less influenced by visual input and exhibits stronger, more stable alpha-band activity, making it ideal for resting-state neural dynamics investigations. Bad channels were identified and removed based on their statistical deviation from the mean signal. Automated cleaning was performed using the *pop_clean_rawdata* function in EEGLAB, which detects and removes flat channels, noisy segments, and power-line noise, thus enhancing reproducibility, saving time, and reducing human bias. This function also provides flexibility through adjustable criteria for detecting various artifacts, and supports interpolation of noisy segments, ensuring minimal data loss. Independent Component Analysis (ICA) was then applied to decompose the EEG signals into independent components, separating neural activity from artifacts such as eye blinks, muscle movements, and cardiac signals. The ICLabel plugin was used to classify components, and non-brain-related components were excluded. Each component’s topography and time series were visually inspected to confirm accurate classification and removal of artifacts. For any detected bad channels, the spherical spline interpolation method was employed to reconstruct missing or corrupted data, preserving spatial consistency. Una cosa che è sempre utile inserire sono media e deviazione del numero di elettrodi interpolati, IC rimosse. Infine è sempre utile dichiarare quanti minuti sono rimasti di segnale dopo la pulizia.

### Source Reconstruction

Source reconstruction was performed using Brainstorm, an open-source toolbox for EEG and MEG analysis. At first, MRIs were segmented using CAT12 toolbox (Gaser et al., 2024). The cortical surface was downsampled to 15002 vertices. Successively, inner skull, outer skull, and scalp meshes were discretized to 1922 vertices. We applied a realistic head model namely, the Boundary Element Models (BEM) using OpenMEEG (Gramfort et al., 2010; Kybic et al., 2005). The inverse solution was estimated using the weighted Minimum Norm Estimation (wMNE) with dipoles orientation constrained normal to surface, using default Brainstorm parameters setting.

Reconstructed source activity was subsequently downsampled to the Desikan-Killiany anatomical atlas by averaging vertices time-series of each region of interest.

### Fluidity

Fluidity, which quantifies the range of functional patterns spontaneously explored by a network, is defined as the variance of the upper triangle of the FCD matrix. To assess connectivity, we calculated the Phase Locking Value (PLV), defined as:

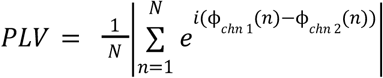

where N is the number of time points within the window, and θ(t)\theta(t)θ(t) represents the phase difference between the two signals at time t. By focusing on phase synchronization, we assumed that stronger synchronization correlates with more communication between brain regions. The PLV was calculated in the canonical frequency bands: theta (4-8 Hz), alpha (8-14 Hz), beta (14-30 Hz), and gamma (30-40 Hz).

The PLV was computed using a sliding window, whose length varied depending on the frequency band analyzed, allowing for the assessment of connectivity changes over time, as in Breyton et al. (Breyton et al., 2024):

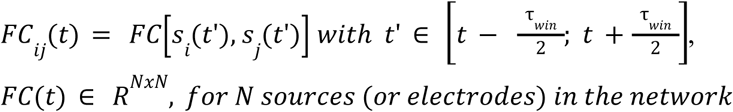

where FC(t) reflects the time-varying connectivity between different brain regions.

Subsequently, Dynamic functional connectivity (FCD) was then calculated by correlating the upper triangular part of PLV matrices from consecutive windows, capturing the temporal evolution of inter-regional synchronization.

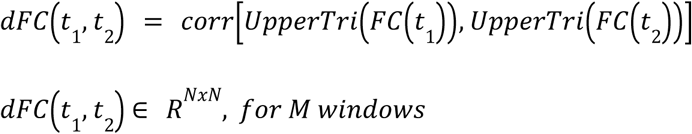

The fluidity was then defined as the variance of the upper triangle of the FCD matrix, after removal of the overlapping windows (Breyton et al., 2024):

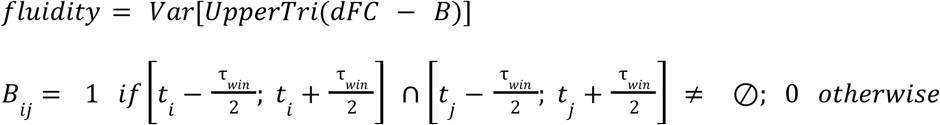

with the diagonal offset to compute *fluidity* given by the following formula:

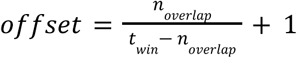

This approach yielded a time-resolved functional connectivity matrix, capturing the dynamics of the inter-regional synchronization. By analyzing the variance of PLV across these sliding windows, the temporal variability of functional connectivity was understood as a proxy of the ability of the brain to dynamically adapt, with higher variance corresponding to a stronger tendency to spontaneous reorganization.

### Statistical Analysis

We computed the average fluidity across all matrices for each subject in both the Alzheimer’s disease (AD) and Control groups. To assess whether the observed difference in the group means was statistically significant, we employed a non-parametric permutation test. Specifically, we randomly shuffled the subject labels across groups while preserving the number of participants in each group. At each iteration (n = 20,000), we computed the absolute difference in means. This generated a null distribution of differences. The observed difference between the original group means was then compared to the null distribution. The p-value was estimated as the proportion of permuted differences that exceeded the observed one. This approach does not assume the normality of the underlying distributions and provides a robust, distribution-free method for significance testing.

Correlations were assessed using Pearson’s correlation. A significance level of 0.05 has been applied.

### Multilinear Model

To test the hypothesis that fluidity in the beta band may serve as a biomarker for Alzheimer’s disease (AD), and to investigate whether it is associated with clinical outcomes, we implemented a Multiple linear regression model to predict, cross-sectionally, cognitive performance as measured by the Mini-Mental State Examination (MMSE) score. In addition to beta-band fluidity, we included three nuisance covariates—age, sex, and total tau (tTau) levels—to control for their potential confounding influence on MMSE. All predictor variables were standardized using z-scores prior to model fitting to facilitate interpretation and ensure comparability of regression coefficients.

The model was validated through a leave-one-out cross-validation (LOOCV) scheme. In each iteration, one subject was held out as the test case while the model was trained on the remaining participants, allowing us to estimate prediction error and model generalizability minimizing the risk of overfitting.

The stability of model coefficients and explained variance was assessed across iterations, and multicollinearity was examined using the Variance Inflation Factor (VIF).

## Data Availability

All data produced in the present study are available upon reasonable request to the authors

## Acknowledgments

This project has received funding from the European Union’s Horizon Europe research and innovation programme under grant agreement No 101147319 (EBRAINS 2.0). We also acknowledge a contribution from the Italian National Recovery and Resilience Plan (NRRP), M4C2, funded by the European Union—NextGenerationEU (Project IR0000011, CUP B51E22000150006, EBRAINS-Italy).

## Bibliography

1. Albert, M.S., DeKosky, S.T., Dickson, D., Dubois, B., Feldman, H.H., Fox, N.C., Gamst, A., Holtzman, D.M., Jagust, W.J., Petersen, R.C., Snyder, P.J., Carrillo, M.C., Thies, B., Phelps, C.H., 2011. The diagnosis of mild cognitive impairment due to Alzheimer’s disease: recommendations from the National Institute on Aging-Alzheimer’s Association workgroups on diagnostic guidelines for Alzheimer’s disease. Alzheimers Dement. J. Alzheimers Assoc. 7, 270–279. 10.1016/j.jalz.2011.03.008

2. Allain, P., Etcharry-Bouyx, F., Verny, C., 2013. Executive functions in clinical and preclinical Alzheimer’s disease. Rev. Neurol. (Paris), Démences : nouveaux concepts, nouveaux enjeux / Dementia: new concepts, new goals 169, 695–708. 10.1016/j.neurol.2013.07.020

3. Babiloni, C., Triggiani, A.I., Lizio, R., Cordone, S., Tattoli, G., Bevilacqua, V., Soricelli, A., Ferri, R., Nobili, F., Gesualdo, L., Millán-Calenti, J.C., Buján, A., Tortelli, R., Cardinali, V., Barulli, M.R., Giannini, A., Spagnolo, P., Armenise, S., Buenza, G., Scianatico, G., Logroscino, G., Frisoni, G.B., del Percio, C., 2016. Classification of Single Normal and Alzheimer’s Disease Individuals from Cortical Sources of Resting State EEG Rhythms. Front. Neurosci. 10. 10.3389/fnins.2016.00047

4. Bastos, A.M., Schoffelen, J.-M., 2015. A Tutorial Review of Functional Connectivity Analysis Methods and Their Interpretational Pitfalls. Front. Syst. Neurosci. 9, 175. 10.3389/fnsys.2015.00175

5. Battaglia, D., Boudou, T., Hansen, E.C.A., Lombardo, D., Chettouf, S., Daffertshofer, A., McIntosh, A.R., Zimmermann, J., Ritter, P., Jirsa, V., 2020. Dynamic Functional Connectivity between order and randomness and its evolution across the human adult lifespan. NeuroImage 222, 117156. 10.1016/j.neuroimage.2020.117156

6. Belleville, S., 2010. S3-02-05: Working memory and control of attention in persons with Alzheimer’s disease and mild cognitive impairment. Alzheimers Dement. 6, S122–S122. 10.1016/j.jalz.2010.05.380

7. Benchenane, K., Tiesinga, P.H., Battaglia, F.P., 2011. Oscillations in the prefrontal cortex: a gateway to memory and attention. Curr. Opin. Neurobiol., Behavioural and cognitive neuroscience 21, 475–485. 10.1016/j.conb.2011.01.004

8. Bierer, L.M., Hof, P.R., Purohit, D.P., Carlin, L., Schmeidler, J., Davis, K.L., Perl, D.P., 1995. Neocortical Neurofibrillary Tangles Correlate With Dementia Severity in Alzheimer’s Disease. Arch. Neurol. 52, 81–88. 10.1001/archneur.1995.00540250089017

9. Botto, R., Callai, N., Cermelli, A., Causarano, L., Rainero, I., 2022. Anxiety and depression in Alzheimer’s disease: a systematic review of pathogenetic mechanisms and relation to cognitive decline. Neurol. Sci. 43, 4107–4124. 10.1007/s10072-022-06068-x

10. Breyton, M., Fousek, J., Rabuffo, G., Sorrentino, P., Kusch, L., Massimini, M., Petkoski, S., Jirsa, V., 2024. Spatiotemporal brain complexity quantifies consciousness outside of perturbation paradigms. eLife 13. 10.7554/eLife.98920.1

11. Buckley, R.F., Saling, M.M., Frommann, I., Wolfsgruber, S., Wagner, M., 2015. Subjective Cognitive Decline from a Phenomenological Perspective: A Review of the Qualitative Literature. J. Alzheimers Dis. JAD 48 Suppl 1, S125–140. 10.3233/JAD-150095

12. Cacciamani, F., Houot, M., Gagliardi, G., Dubois, B., Sikkes, S., Sánchez-Benavides, G., Denicolò, E., Molinuevo, J.L., Vannini, P., Epelbaum, S., 2021. Awareness of Cognitive Decline in Patients With Alzheimer’s Disease: A Systematic Review and Meta-Analysis. Front. Aging Neurosci. 13, 697234. 10.3389/fnagi.2021.697234

13. Canuet, L., Tellado, I., Couceiro, V., Fraile, C., Fernandez-Novoa, L., Ishii, R., Takeda, M., Cacabelos, R., 2012. Resting-State Network Disruption and APOE Genotype in Alzheimer’s Disease: A lagged Functional Connectivity Study. PLOS ONE 7, e46289. 10.1371/journal.pone.0046289

14. Cao, J., Zhao, Y., Shan, X., Wei, H., Guo, Y., Chen, L., Erkoyuncu, J.A., Sarrigiannis, P.G., 2022. Brain functional and effective connectivity based on electroencephalography recordings: A review. Hum. Brain Mapp. 43, 860–879. 10.1002/hbm.25683

15. Christoff, K., Ream, J.M., Geddes, L.P.T., Gabrieli, J.D.E., 2003. Evaluating Self-Generated Information: Anterior Prefrontal Contributions to Human Cognition. Behav. Neurosci. 117, 1161–1168. 10.1037/0735-7044.117.6.1161

16. Cipriano, L., Minino, R., Liparoti, M., Polverino, A., Romano, A., Bonavita, S., Pirozzi, M.A., Quarantelli, M., Jirsa, V., Sorrentino, G., Sorrentino, P., Troisi Lopez, E., 2024. Flexibility of brain dynamics is increased and predicts clinical impairment in relapsing-remitting but not in secondary progressive multiple sclerosis. Brain Commun. 6, fcae112. 10.1093/braincomms/fcae112

17. Cohen, M.X., 2014. Analyzing Neural Time Series Data: Theory and Practice. The MIT Press. 10.7551/mitpress/9609.001.0001

18. Cummings, J.L., Raman, R., Ernstrom, K., Salmon, D., Ferris, S.H., Group, for the A.D.C.S., 2006. ADCS Prevention Instrument Project: Behavioral Measures in Primary Prevention Trials. Alzheimer Dis. Assoc. Disord. 20, S147. 10.1097/01.wad.0000213872.17429.0f

19. Dauwels, J., Vialatte, F., Cichocki, A., n.d. Diagnosis of Alzheimers Disease from EEG Signals: Where Are We Standing? Curr. Alzheimer Res. 7, 487–505. 10.2174/156720510792231720

20. Delbeuck, X., Collette, F., Van der Linden, M., 2007. Is Alzheimer’s disease a disconnection syndrome?: Evidence from a crossmodal audio-visual illusory experiment. Neuropsychologia 45, 3315–3323. 10.1016/j.neuropsychologia.2007.05.001

21. Dubois, B., Hampel, H., Feldman, H.H., Scheltens, P., Aisen, P., Andrieu, S., Bakardjian, H., Benali, H., Bertram, L., Blennow, K., Broich, K., Cavedo, E., Crutch, S., Dartigues, J.-F., Duyckaerts, C., Epelbaum, S., Frisoni, G.B., Gauthier, S., Genthon, R., Gouw, A.A., Habert, M.-O., Holtzman, D.M., Kivipelto, M., Lista, S., Molinuevo, J.-L., O’Bryant, S.E., Rabinovici, G.D., Rowe, C., Salloway, S., Schneider, L.S., Sperling, R., Teichmann, M., Carrillo, M.C., Cummings, J., Jack, C.R., Proceedings of the Meeting of the International Working Group (IWG) and the American Alzheimer’s Association on “The Preclinical State of AD”; July 23, 2015; Washington DC, USA, 2016. Preclinical Alzheimer’s disease: Definition, natural history, and diagnostic criteria. Alzheimers Dement. J. Alzheimers Assoc. 12, 292–323. 10.1016/j.jalz.2016.02.002

22. Duncan, J., 2005. Frontal Lobe Function and General Intelligence: Why it Matters. Cortex 41, 215–217. 10.1016/S0010-9452(08)70896-7

23. Duncan, J., Burgess, P., Emslie, H., 1995. Fluid intelligence after frontal lobe lesions. Neuropsychologia 33, 261–268. 10.1016/0028-3932(94)00124-8

24. Elsiddig, A.A.I., Grosu, C., Ferrer Soler, C., Scheffler, M., Cotta Ramusino, M., Trombella, S., Gold, G., Boccardi, M., Frisoni, G.B., 2018. [MRI of medial-temporal atrophy as a biomarker for Alzheimer’s disease]. Rev. Med. Suisse 14, 1716–1720.

25. Engel, A.K., Fries, P., 2010. Beta-band oscillations — signalling the status quo? Curr. Opin. Neurobiol., Cognitive neuroscience 20, 156–165. 10.1016/j.conb.2010.02.015

26. Engels, M.M., Stam, C.J., van der Flier, W.M., Scheltens, P., de Waal, H., van Straaten, E.C., 2015. Declining functional connectivity and changing hub locations in Alzheimer’s disease: an EEG study. BMC Neurol. 15, 145. 10.1186/s12883-015-0400-7

27. Folstein, M.F., Folstein, S.E., McHugh, P.R., 1975. “Mini-mental state”. A practical method for grading the cognitive state of patients for the clinician. J. Psychiatr. Res. 12, 189–198. 10.1016/0022-3956(75)90026-6

28. Fornito, A., Zalesky, A., Breakspear, M., 2015. The connectomics of brain disorders. Nat. Rev. Neurosci. 16, 159–172. 10.1038/nrn3901

29. Friston, K.J., 2011. Functional and effective connectivity: a review. Brain Connect. 1, 13–36. 10.1089/brain.2011.0008

30. Fukutani, Y., Cairns, N.J., Shiozawa, M., Sasaki, K., Sudo, S., Isaki, K., Lantos, P.L., 2000. Neuronal loss and neurofibrillary degeneration in the hippocampal cortex in late-onset sporadic Alzheimer’s disease. Psychiatry Clin. Neurosci. 54, 523–529. 10.1046/j.1440-1819.2000.00747.x

31. Gallego-Rudolf, J., Wiesman, A.I., Pichet Binette, A., Villeneuve, S., Baillet, S., 2024. Synergistic association of Aβ and tau pathology with cortical neurophysiology and cognitive decline in asymptomatic older adults. Nat. Neurosci. 27, 2130–2137. 10.1038/s41593-024-01763-8

32. Gaser, C., Dahnke, R., Thompson, P.M., Kurth, F., Luders, E., The Alzheimer’s Disease Neuroimaging Initiative, null, 2024. CAT: a computational anatomy toolbox for the analysis of structural MRI data. GigaScience 13, giae049. 10.1093/gigascience/giae049

33. Giebel, C.M., Sutcliffe, C., Stolt, M., Karlsson, S., Renom-Guiteras, A., Soto, M., Verbeek, H., Zabalegui, A., Challis, D., 2014. Deterioration of basic activities of daily living and their impact on quality of life across different cognitive stages of dementia: a European study. Int. Psychogeriatr. 26, 1283–1293. 10.1017/S1041610214000775

34. Gramfort, A., Papadopoulo, T., Olivi, E., Clerc, M., 2010. OpenMEEG: opensource software for quasistatic bioelectromagnetics. Biomed. Eng. OnLine 9, 45. 10.1186/1475-925X-9-45

35. Hampel, H., Hardy, J., Blennow, K., Chen, C., Perry, G., Kim, S.H., Villemagne, V.L., Aisen, P., Vendruscolo, M., Iwatsubo, T., Masters, C.L., Cho, M., Lannfelt, L., Cummings, J.L., Vergallo, A., 2021. The Amyloid-β Pathway in Alzheimer’s Disease. Mol. Psychiatry 26, 5481–5503. 10.1038/s41380-021-01249-0

36. Hampel, H., Toschi, N., Babiloni, C., Baldacci, F., Black, K.L., Bokde, A.L.W., Bun, R.S., Cacciola, F., Cavedo, E., Chiesa, P.A., Colliot, O., Coman, C.-M., Dubois, B., Duggento, A., Durrleman, S., Ferretti, M.-T., George, N., Genthon, R., Habert, M.-O., Herholz, K., Koronyo, Y., Koronyo-Hamaoui, M., Lamari, F., Langevin, T., Lehéricy, S., Lorenceau, J., Neri, C., Nisticò, R., Nyasse-Messene, F., Ritchie, C., Rossi, S., Santarnecchi, E., Sporns, O., Verdooner, S.R., Vergallo, A., Villain, N., Younesi, E., Garaci, F., Lista, S., 2018. Revolution of Alzheimer Precision Neurology. Passageway of Systems Biology and Neurophysiology. J. Alzheimer’s Dis. 64, S47–S105. 10.3233/JAD-179932

37. Hansen, E.C.A., Battaglia, D., Spiegler, A., Deco, G., Jirsa, V.K., 2015. Functional connectivity dynamics: Modeling the switching behavior of the resting state. NeuroImage 105, 525–535. 10.1016/j.neuroimage.2014.11.001

38. Hata, M., Kazui, H., Tanaka, T., Ishii, R., Canuet, L., Pascual-Marqui, R.D., Aoki, Y., Ikeda, S., Kanemoto, H., Yoshiyama, K., Iwase, M., Takeda, M., 2016. Functional connectivity assessed by resting state EEG correlates with cognitive decline of Alzheimer’s disease - An eLORETA study. Clin. Neurophysiol. Off. J. Int. Fed. Clin. Neurophysiol. 127, 1269–1278. 10.1016/j.clinph.2015.10.030

39. Heitmann, S., Breakspear, M., 2018. Putting the “dynamic” back into dynamic functional connectivity. Netw. Neurosci. 2, 150. 10.1162/netn_a_00041

40. Herweg, N.A., Solomon, E.A., Kahana, M.J., 2020. Theta Oscillations in Human Memory. Trends Cogn. Sci. 24, 208–227. 10.1016/j.tics.2019.12.006

41. Hutchison, R.M., Womelsdorf, T., Gati, J.S., Everling, S., Menon, R.S., 2013. Resting-state networks show dynamic functional connectivity in awake humans and anesthetized macaques: Dynamic Functional Connectivity. Hum. Brain Mapp. 34, 2154–2177. 10.1002/hbm.22058

42. Ibanez, A., Kringelbach, M.L., Deco, G., 2024. A synergetic turn in cognitive neuroscience of brain diseases. Trends Cogn. Sci. 28, 319–338. 10.1016/j.tics.2023.12.006

43. Ismail, Z., Rajji, T.K., Shulman, K.I., 2010. Brief cognitive screening instruments: an update. Int. J. Geriatr. Psychiatry 25, 111–120. 10.1002/gps.2306

44. Jacini, F., Sorrentino, P., Lardone, A., Rucco, R., Baselice, F., Cavaliere, C., Aiello, M., Orsini, M., Iavarone, A., Manzo, V., Carotenuto, A., Granata, C., Hillebrand, A., Sorrentino, G., 2018. Amnestic Mild Cognitive Impairment Is Associated With Frequency-Specific Brain Network Alterations in Temporal Poles. Front. Aging Neurosci. 10, 400. 10.3389/fnagi.2018.00400

45. Jack Jr., C.R., Andrews, J.S., Beach, T.G., Buracchio, T., Dunn, B., Graf, A., Hansson, O., Ho, C., Jagust, W., McDade, E., Molinuevo, J.L., Okonkwo, O.C., Pani, L., Rafii, M.S., Scheltens, P., Siemers, E., Snyder, H.M., Sperling, R., Teunissen, C.E., Carrillo, M.C., 2024. Revised criteria for diagnosis and staging of Alzheimer’s disease: Alzheimer’s Association Workgroup. Alzheimers Dement. 20, 5143–5169. 10.1002/alz.13859

46. Jafari, Z., Kolb, B.E., Mohajerani, M.H., 2020. Neural oscillations and brain stimulation in Alzheimer’s disease. Prog. Neurobiol. 194, 101878. 10.1016/j.pneurobio.2020.101878

47. Jahn, H., 2013. Memory loss in Alzheimer’s disease. Dialogues Clin. Neurosci. 15, 445–454. 10.31887/DCNS.2013.15.4/hjahn

48. Jansen, W.J., Ossenkoppele, R., Knol, D.L., Tijms, B.M., Scheltens, P., Verhey, F.R.J., Visser, P.J., and the Amyloid Biomarker Study Group, 2015. Prevalence of Cerebral Amyloid Pathology in Persons Without Dementia: A Meta-analysis. JAMA 313, 1924–1938. 10.1001/jama.2015.4668

49. Jeong, J., 2004. EEG dynamics in patients with Alzheimer’s disease. Clin. Neurophysiol. Off. J. Int. Fed. Clin. Neurophysiol. 115, 1490–1505. 10.1016/j.clinph.2004.01.001

50. Kabbara, A., Eid, H., El Falou, W., Khalil, M., Wendling, F., Hassan, M., 2018. Reduced integration and improved segregation of functional brain networks in Alzheimer’s disease. J. Neural Eng. 15, 026023. 10.1088/1741-2552/aaaa76

51. Kehoe, E.G., McNulty, J.P., Mullins, P.G., Bokde, A.L.W., 2014. Advances in MRI biomarkers for the diagnosis of Alzheimer’s disease. Biomark. Med. 8, 1151–1169. 10.2217/bmm.14.42

52. Kurkinen, M., Fułek, M., Fułek, K., Beszłej, J.A., Kurpas, D., Leszek, J., 2023. The Amyloid Cascade Hypothesis in Alzheimer’s Disease: Should We Change Our Thinking? Biomolecules 13, 453. 10.3390/biom13030453

53. Kybic, J., Clerc, M., Abboud, T., Faugeras, O., Keriven, R., Papadopoulo, T., 2005. A common formalism for the integral formulations of the forward EEG problem. IEEE Trans. Med. Imaging 24, 12–28. 10.1109/tmi.2004.837363

54. Lachaux, J., Rodriguez, E., Martinerie, J., Varela, F.J., 1999. Measuring phase synchrony in brain signals. Hum. Brain Mapp. 8, 194–208. 10.1002/(SICI)1097-0193(1999)8:4<194::AID-HBM4>3.0.CO;2-C

55. Lee, K.H., Choi, Y.Y., Gray, J.R., Cho, S.H., Chae, J.-H., Lee, S., Kim, K., 2006. Neural correlates of superior intelligence: Stronger recruitment of posterior parietal cortex. NeuroImage 29, 578–586. 10.1016/j.neuroimage.2005.07.036

56. Liparoti, M., Cipriano, L., Troisi Lopez, E., Polverino, A., Minino, R., Sarno, L., Sorrentino, G., Lucidi, F., Sorrentino, P., 2024. Brain flexibility increases during the peri-ovulatory phase as compared to early follicular phase of the menstrual cycle. Sci. Rep. 14, 1976. 10.1038/s41598-023-49588-y

57. Maresova, P., Hruska, J., Klimova, B., Barakovic, S., Krejcar, O., 2020. Activities of Daily Living and Associated Costs in the Most Widespread Neurodegenerative Diseases: A Systematic Review. Clin. Interv. Aging 15, 1841–1862. 10.2147/CIA.S264688

58. Mega, M.S., Cummings, J.L., Fiorello, T., Gornbein, J., 1996. The spectrum of behavioral changes in Alzheimer’s disease. Neurology 46, 130–135. 10.1212/WNL.46.1.130

59. Mielke, M.M., 2018. Sex and Gender Differences in Alzheimer’s Disease Dementia. Psychiatr. Times 35, 14.

60. Monacelli, A.M., Cushman, L.A., Kavcic, V., Duffy, C.J., 2003. Spatial disorientation in Alzheimer’s disease. Neurology 61, 1491–1497. 10.1212/WNL.61.11.1491

61. Morganti, F., Stefanini, S., Riva, G., 2013. From allo-to egocentric spatial ability in early Alzheimer’s disease: a study with virtual reality spatial tasks. Cogn. Neurosci. 4, 171–180. 10.1080/17588928.2013.854762

62. Nobili, F., Arbizu, J., Bouwman, F., Drzezga, A., Agosta, F., Nestor, P., Walker, Z., Boccardi, M., EANM-EAN Task Force for the Prescription of FDG-PET for Dementing Neurodegenerative Disorders, 2018. European Association of Nuclear Medicine and European Academy of Neurology recommendations for the use of brain 18 F-fluorodeoxyglucose positron emission tomography in neurodegenerative cognitive impairment and dementia: Delphi consensus. Eur. J. Neurol. 25, 1201–1217. 10.1111/ene.13728

63. Paitel, E.R., Otteman, C.B.D., Polking, M.C., Licht, H.J., Nielson, K.A., 2025. Functional and effective EEG connectivity patterns in Alzheimer’s disease and mild cognitive impairment: a systematic review. Front. Aging Neurosci. 17, 1496235. 10.3389/fnagi.2025.1496235

64. Palmigiano, A., Geisel, T., Wolf, F., Battaglia, D., 2017. Flexible information routing by transient synchrony. Nat. Neurosci. 20, 1014–1022. 10.1038/nn.4569

65. Palop, J.J., Mucke, L., 2016. Network abnormalities and interneuron dysfunction in Alzheimer disease. Nat. Rev. Neurosci. 17, 777–792. 10.1038/nrn.2016.141

66. Peters-Founshtein, G., Gazit, L., Naveh, T., Domachevsky, L., Korczyn, A.D., Bernstine, H., Shaharabani-Gargir, L., Groshar, D., Marshall, G.A., Arzy, S., 2024. Lost in space(s): Multimodal neuroimaging of disorientation along the Alzheimer’s disease continuum. Hum. Brain Mapp. 45, e26623. 10.1002/hbm.26623

67. Polverino, A., Troisi Lopez, E., Minino, R., Liparoti, M., Romano, A., Trojsi, F., Lucidi, F., Gollo, L., Jirsa, V., Sorrentino, G., Sorrentino, P., 2022. Flexibility of Fast Brain Dynamics and Disease Severity in Amyotrophic Lateral Sclerosis. Neurology 99, e2395–e2405. 10.1212/WNL.0000000000201200

68. Preti, M.G., Bolton, T.A., Van De Ville, D., 2017. The dynamic functional connectome: State-of-the-art and perspectives. NeuroImage, Functional Architecture of the Brain 160, 41–54. 10.1016/j.neuroimage.2016.12.061

69. Romano, A., Troisi Lopez, E., Cipriano, L., Liparoti, M., Minino, R., Polverino, A., Cavaliere, C., Aiello, M., Granata, C., Sorrentino, G., Sorrentino, P., 2023. Topological changes of fast large-scale brain dynamics in mild cognitive impairment predict early memory impairment: a resting-state, source reconstructed, magnetoencephalography study. Neurobiol. Aging 132, 36–46. 10.1016/j.neurobiolaging.2023.08.003

70. Rueda, A.D., Lau, K.M., Saito, N., Harvey, D., Risacher, S.L., Aisen, P.S., Petersen, R.C., Saykin, A.J., Farias, S.T., Initiative, A.D.N., 2015. Self-rated and informant-rated everyday function in comparison to objective markers of Alzheimer’s disease. Alzheimers Dement. 11, 1080–1089. 10.1016/j.jalz.2014.09.002

71. Smailovic, U., Jelic, V., 2019. Neurophysiological Markers of Alzheimer’s Disease: Quantitative EEG Approach. Neurol. Ther. 8, 37–55. 10.1007/s40120-019-00169-0

72. Sorrentino, P., Rucco, R., Baselice, F., De Micco, R., Tessitore, A., Hillebrand, A., Mandolesi, L., Breakspear, M., Gollo, L.L., Sorrentino, G., 2021. Flexible brain dynamics underpins complex behaviours as observed in Parkinson’s disease. Sci. Rep. 11. 10.1038/S41598-021-83425-4

73. Stam, C.J., 2014. Modern network science of neurological disorders. Nat. Rev. Neurosci. 15, 683–695. 10.1038/nrn3801

74. Stoll, F.M., Wilson, C.R.E., Faraut, M.C.M., Vezoli, J., Knoblauch, K., Procyk, E., 2016. The Effects of Cognitive Control and Time on Frontal Beta Oscillations. Cereb. Cortex 26, 1715–1732. 10.1093/cercor/bhv006

75. Tekin, S., Fairbanks, L.A., O’Connor, S., Rosenberg, S., Cummings, J.L., 2001. Activities of Daily Living in Alzheimer’s Disease: Neuropsychiatric, Cognitive, and Medical Illness Influences. Am. J. Geriatr. Psychiatry 9, 81–86. 10.1097/00019442-200102000-00013

76. Uddin, M.S., Ashraf, G.M., Uddin, M.S., Ashraf, G.M., 2018. Introductory Chapter: Alzheimer’s Disease—The Most Common Cause of Dementia, in: Advances in Dementia Research. IntechOpen. 10.5772/intechopen.82196

77. Vecchio, F., Miraglia, F., Marra, C., Quaranta, D., Vita, M.G., Bramanti, P., Rossini, P.M., 2014. Human brain networks in cognitive decline: a graph theoretical analysis of cortical connectivity from EEG data. J. Alzheimers Dis. JAD 41, 113–127. 10.3233/JAD-132087

78. Verma, M., Howard, R.J., 2012. Semantic memory and language dysfunction in early Alzheimer’s disease: a review. Int. J. Geriatr. Psychiatry 27, 1209–1217. 10.1002/gps.3766

79. Watanabe, H., Bagarinao, E., Yokoi, T., Yamaguchi, H., Ishigaki, S., Mausuda, M., Katsuno, M., Sobue, G., 2019. Tau Accumulation and Network Breakdown in Alzheimer’s Disease. Adv. Exp. Med. Biol. 1184, 231–240. 10.1007/978-981-32-9358-8_19

80. Yalçınkaya, B.H., Ziaeemehr, A., Fousek, J., Hashemi, M., Lavanga, M., Solodkin, A., McIntosh, A.R., Jirsa, V.K., Petkoski, S., 2023. Personalized virtual brains of Alzheimer’s Disease link dynamical biomarkers of fMRI with increased local excitability. 10.1101/2023.01.11.23284438

81. Yu, M., Sporns, O., Saykin, A.J., 2021. The human connectome in Alzheimer disease -relationship to biomarkers and genetics. Nat. Rev. Neurol. 17, 545–563. 10.1038/s41582-021-00529-1

82. Zalesky, A., Fornito, A., Cocchi, L., Gollo, L.L., Breakspear, M., 2014. Time-resolved resting-state brain networks. Proc. Natl. Acad. Sci. 111, 10341–10346. 10.1073/pnas.1400181111

83. Zhang, Y., Chen, H., Li, R., Sterling, K., Song, W., 2023. Amyloid β-based therapy for Alzheimer’s disease: challenges, successes and future. Signal Transduct. Target. Ther. 8, 1–26. 10.1038/s41392-023-01484-7

84. Zhao, Q.-F., Tan, Lan, Wang, H.-F., Jiang, T., Tan, M.-S., Tan, Lin, Xu, W., Li, J.-Q., Wang, J., Lai, T.-J., Yu, J.-T., 2016. The prevalence of neuropsychiatric symptoms in Alzheimer’s disease: Systematic review and meta-analysis. J. Affect. Disord. 190, 264–271. 10.1016/j.jad.2015.09.069

