## Supplementary Material for "Brain Fluidity as a Functional Marker of Tau-Related Neurodegeneration in Alzheimer’s Disease"

### Supplementary Materials

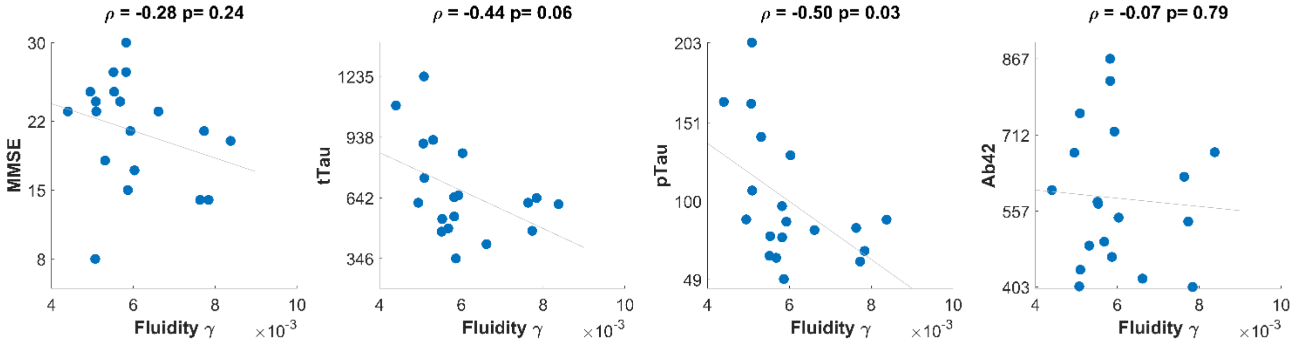


**Fig. S1 Correlations between gamma-band fluidity and clinical/cerebrospinal biomarkers.** Scatter plots show the relationships between gamma-band fluidity and MMSE scores, total tau (tTau), phosphorylated tau (pTau), and amyloid-beta 42 (Aβ42) levels. A significant negative correlation was found between gamma fluidity and pTau levels (ρ=-0.50, p = 0.03).

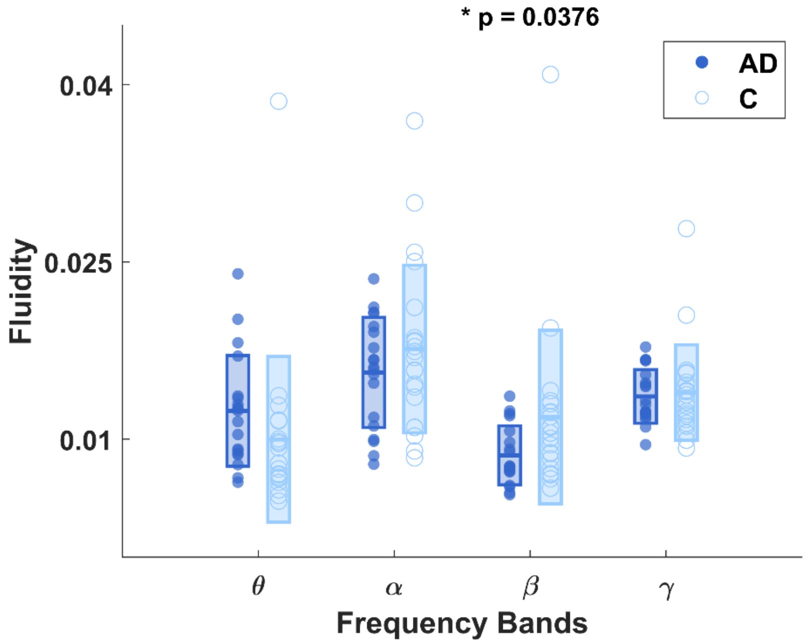


**Fig. S2 Group differences in fluidity across frequency bands at the sensor level**. Distribution of individual fluidity values in Alzheimer's disease (AD) patients and healthy controls across theta (4–8 Hz), alpha (8–14 Hz), beta (14–30 Hz), and gamma (30–40 Hz) frequency bands. AD patients exhibited increased fluidity in the theta band and significantly reduced fluidity in the beta band (p = 0.0376) compared to controls. This pattern is consistent with the results obtained from the source-reconstructed data.
